# Before AI tells patients what to eat, it needs to know where they live: a cross-sectional ecological study of diabetes and food access in California and Michigan census tracts, with a spotlight on Metro Detroit and Metro Los Angeles

**DOI:** 10.64898/2026.11.03.26367208

**Authors:** Sri Sudheera Chitipolu

## Abstract

**Background:** AI chatbots now give people with diabetes diet advice, but whether those foods can be bought near home is rarely checked.

**Methods:** In a cross-sectional ecological study of 11,974 California and Michigan census tracts, I linked diabetes prevalence (CDC PLACES 2025) with two USDA food-access maps: all SNAP-authorized stores (2025) and large grocery stores only (2019). I identified double-burden tracts (high diabetes and low income/low access) and hidden food deserts (low access to large grocery stores, but not to any SNAP store). In a pilot, ChatGPT, Gemini, and Claude generated meal plans for the same person across 20 high-diabetes tracts in Metro Los Angeles and Metro Detroit; I scored whether advice was healthy and whether foods were sold at nearby store types.

**Results:** Double-burden tracts were few (71 in California, 19 in Michigan), almost none in the two metros. Food-desert tracts had a 1.5 percentage-point higher diabetes prevalence (95% CI, 1.0–2.0). Counting all SNAP stores hid 567 Californian and 329 Michigan food deserts; in Metro Detroit, all 97 disappeared, including 51 high-diabetes tracts with about 19,500 adults with diabetes. Chatbot advice was healthy, but in hidden tracts 30% of recommended foods were not reliably sold nearby; the chatbots barely adjusted. Of 55 places they named, 13% had errors, and most were over a mile away.

**Conclusions:** The current USDA map obscures food deserts, especially in Detroit, and AI diet advice barely adjusts for them. AI nutrition tools for diabetes should include a place-aware check built on public food-access data.

## 1. Introduction

Diabetes is not shaped only by medicine and doctor visits. It is also shaped by where people live and what they can afford. Social and environmental conditions, such as income, housing, and neighborhood resources, are estimated to account for 50% to 60% of health outcomes, and the American Diabetes Association identifies food insecurity and limited food access as factors associated with higher diabetes risk and poorer outcomes (Hill-Briggs et al., 2021). Food insecurity is common among people with diabetes: 16% of US adults with diabetes report it, compared with 9% of adults without diabetes (Levi et al., 2023). The barriers people most often describe are practical ones: lack of transportation and no nearby grocery store that sells healthy food (Levi et al., 2023).

However, having fewer nearby stores does not, on its own, explain why some neighborhoods have higher rates of diabetes. A large multi-cohort study found that people in more disadvantaged neighborhoods had a higher risk of type 2 diabetes. Still, the availability of supermarkets and fast-food restaurants explained little or none of this difference (Thorpe et al., 2022). A meta-analysis found no consistent association between the food environment and type 2 diabetes, and noted that studies measured food access in many different ways (den Braver et al., 2018). In Detroit, living in a USDA low-income, low-access tract was not associated with diabetes in pregnancy after adjustment (McClain et al., 2026), even though access in the city has long been unequal: two decades ago, the poorest predominantly African American neighborhoods were on average 1.1 miles farther from a chain supermarket than the poorest White neighborhoods, and 23% to 28% of their households had no car (Zenk et al., 2005). The practical question, then, is not only why diabetes rates are higher in some places, but also where help should go first. Food-is-medicine programs and health insurers operate on limited budgets, and neighborhoods where high diabetes burden and poor food access co-occur are a natural place to start. That information is not readily available today.

Recent studies have begun mapping diabetes at smaller scales, but they still leave gaps. A county-level machine-learning study found poverty and food insecurity to be leading contributors to diabetes prevalence. Still, county averages can mask substantial differences among neighborhoods, and the model performed poorly in regions it had not been trained on (Yahaya et al., 2026, preprint). A study of 5,420 Southern California census tracts found that diagnosed diabetes prevalence ranged from 1.4% to 33.6% between neighborhoods; however, its explanatory measures came from the same Centers for Disease Control and Prevention (CDC) estimation system as the outcome, and it included no direct measure of distance to food stores (Descarpentrie et al., 2025). In New York City, measures drawn from independent federal sources accounted for 67% of the differences in diabetes prevalence between tracts, while USDA low food access accounted for only 2% (Tanner et al., 2024). The size of the area also matters. In a 2026 national study, the link between stores and diabetes pointed in one direction when counties were compared and in the opposite direction when census tracts were compared (Bhagavathula & Williams, 2026). So does the type of store. Across more than 19,000 urban census tracts, living near more dollar stores was associated with higher diabetes prevalence, and living near more grocery stores was associated with lower prevalence (Bruellman et al., 2026). Prior work has also called for testing other definitions of low food access (McClain et al., 2026). This matters because a neighborhood can appear to have access when its only nearby store is a convenience or dollar store. To my knowledge, no study has compared tract-level diabetes and objectively measured food access across California and Michigan, or between their largest metropolitan areas, using current national data.

Meanwhile, chatbots are being tested to help people manage diabetes. In a review of 25 studies, most chatbots provided advice on diet, exercise, blood sugar checks, and medications, but the evidence remained weak (Wu et al., 2024). Newer AI tools, called large language models, now also give diet advice. When three large language models created meal plans for 24 simulated adults with type 2 diabetes, none fully followed the nutrition care process; all exceeded fat and saturated-fat targets, and dietitian reviewers noted that some models did not assess access to food (Bayram et al., 2025). Chatbots are already part of food-is-medicine programs: a bilingual chatbot supported a produce-prescription trial for pregnant women in Houston, without considering users’ access to stores, transportation, or cost (Macias-Navarro et al., 2026). Professional nutrition organizations now call for standards on how AI is designed, deployed, and evaluated, but their guidance does not address food access (Murofushi et al., 2026). A chatbot may recommend fresh salmon and leafy greens to someone whose only nearby store is a dollar store. A few studies have begun to include location and budget in their prompts. For a simulated patient with breast cancer, two chatbots adjusted their grocery lists to reflect culture and budget, and one displayed a map of nearby stores for a given ZIP code. However, the authors found that location barely changed the foods the chatbots recommended, and they did not check whether those foods were sold nearby (Logan et al., 2025). To my knowledge, no study has evaluated AI-generated diabetes diet advice against the kinds of stores near where patients live.

This study had three aims. First, to find census tracts in California and Michigan where high diagnosed diabetes prevalence and low-income, low-access food environments occur together (a ‘double burden’), and to estimate how many adults live in them. I compare the two states overall, urban tracts with urban tracts, rural tracts with rural tracts, and their largest metropolitan areas, Metro Los Angeles and Metro Detroit. Second, to test whether this picture changes when food access counts every store that accepts SNAP benefits (2025) rather than supermarkets only (2019). Third, to pilot a place-aware audit: when a resident of these neighborhoods asks an AI chatbot for meal advice, does it recommend foods that can be bought nearby? The study is descriptive. It shows where problems overlap, not what causes them.

## 2. Methods

### 2.1 Study design and setting

This was a cross-sectional ecological study: I examined neighborhoods at a single point in time, not individuals. The unit of analysis was the 2020 census tract. I included all census tracts in California and Michigan and compared them at two levels: (1) state with state, overall and separately for urban and rural tracts as classified by USDA; and (2) metropolitan area with metropolitan area, using the official Office of Management and Budget delineations of July 2023 (Office of Management and Budget, 2023) of the Los Angeles–Long Beach–Anaheim metropolitan statistical area (Los Angeles and Orange counties) and the Detroit–Warren–Dearborn metropolitan statistical area (Wayne, Oakland, Macomb, Livingston, St. Clair and Lapeer counties). Comparing like with like at both levels avoids comparing a whole state with a single city.

### 2.2 Data sources

Diabetes. Table 1 summarizes all data sources and how I used each one. Tract-level diagnosed diabetes prevalence among adults came from CDC PLACES 2025. PLACES does not count every person with diabetes; it estimates prevalence using multilevel regression and poststratification of Behavioral Risk Factor Surveillance System survey data, based on individual age, sex, race/ethnicity, and education, and county-level poverty, applied to 2020 census block populations (Greenlund et al., 2022; CDC, 2025). I used the crude prevalence estimate, the only version provided at the tract level, and its 95% confidence interval. Because crude rates are not adjusted for age, differences between the two states partly reflect differences in age structure; I therefore compared tracts within each state and included the share of residents aged 65 and older in sensitivity models (USDA SRAM file: residents aged 65+ divided by 2020 tract population).

**Table 1.**
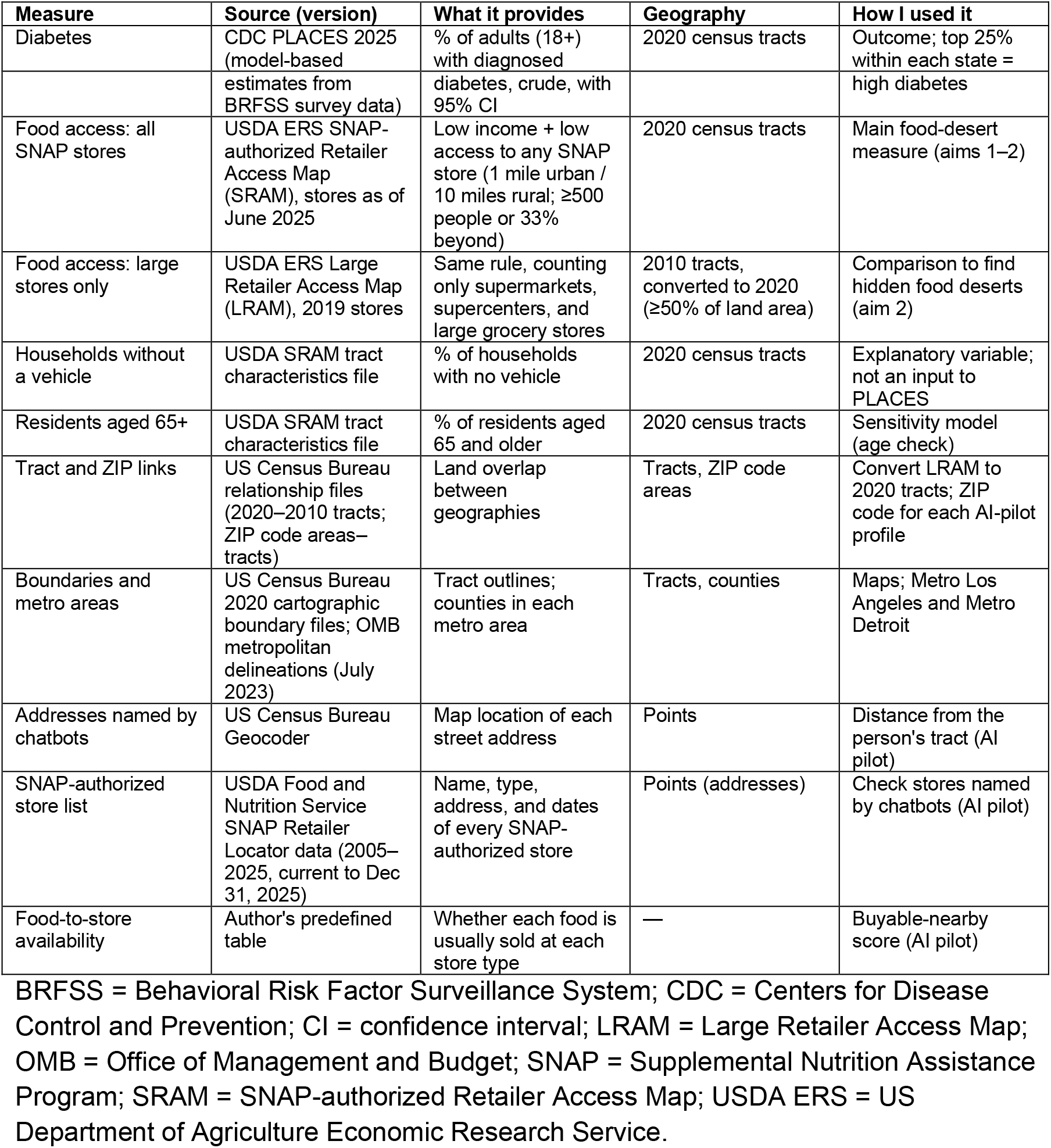
Data sources and how each was used.

| Measure | Source (version) | What it provides | Geography | How I used it |
|---|---|---|---|---|
| Diabetes | CDC PLACES 2025 (model-based estimates from BRFSS survey data) | % of adults (18+) with diagnosed diabetes, crude, with 95% CI | 2020 census tracts | Outcome; top 25% within each state = high diabetes |
| Food access: all SNAP stores | USDA ERS SNAP-authorized Retailer Access Map (SRAM), stores as of June 2025 | Low income + low access to any SNAP store (1 mile urban / 10 miles rural; ≥500 people or 33% beyond) | 2020 census tracts | Main food-desert measure (aims 1–2) |
| Food access: large stores only | USDA ERS Large Retailer Access Map (LRAM), 2019 stores | Same rule, counting only supermarkets, supercenters, and large grocery stores | 2010 tracts, converted to 2020 (≥50% of land area) | Comparison to find hidden food deserts (aim 2) |
| Households without a vehicle | USDA SRAM tract characteristics file | % of households with no vehicle | 2020 census tracts | Explanatory variable; not an input to PLACES |
| Residents aged 65+ | USDA SRAM tract characteristics file | % of residents aged 65 and older | 2020 census tracts | Sensitivity model (age check) |
| Tract and ZIP links | US Census Bureau relationship files (2020–2010 tracts; ZIP code areas–tracts) | Land overlap between geographies | Tracts, ZIP code areas | Convert LRAM to 2020 tracts; ZIP code for each AI-pilot profile |
| Boundaries and metro areas | US Census Bureau 2020 cartographic boundary files; OMB metropolitan delineations (July 2023) | Tract outlines; counties in each metro area | Tracts, counties | Maps; Metro Los Angeles and Metro Detroit |
| Addresses named by chatbots | US Census Bureau Geocoder | Map location of each street address | Points | Distance from the person's tract (AI pilot) |
| SNAP-authorized store list | USDA Food and Nutrition Service SNAP Retailer Locator data (2005–2025, current to Dec 31, 2025) | Name, type, address, and dates of every SNAP-authorized store | Points (addresses) | Check stores named by chatbots (AI pilot) |
| Food-to-store availability | Author's predefined table | Whether each food is usually sold at each store type | — | Buyable-nearby score (AI pilot) |

BRFSS = Behavioral Risk Factor Surveillance System; CDC = Centers for Disease Control and Prevention; CI = confidence interval; LRAM = Large Retailer Access Map; OMB = Office of Management and Budget; SNAP = Supplemental Nutrition Assistance Program; SRAM = SNAP-authorized Retailer Access Map; USDA ERS = US Department of Agriculture Economic Research Service.

Food access. Food access came from the USDA Economic Research Service Food Access Research Atlas, which provides two measures (USDA ERS, 2026). The SNAP-authorized Retailer Access Map (SRAM) measures distance to any store that accepts SNAP benefits, including supermarkets, small grocery stores, convenience stores, and dollar stores, using retailers authorized as of June 2025 and 2020 census tracts. The Large Retailer Access Map (LRAM) measures distance only to supermarkets, supercenters, and large grocery stores, using 2019 store data and 2010 census tracts, which I converted to 2020 tracts using the Census Bureau tract relationship file (U.S. Census Bureau, 2026). In both measures, a tract is classified as low income using New Markets Tax Credit criteria and as low access using distance thresholds of 1 mile in urban areas and 10 miles in rural areas; a tract is low access when at least 500 people, or 33% of its population, live beyond the threshold (USDA ERS, 2026).

Households without a vehicle. The percentage of households without a vehicle came from the USDA SRAM General Tract Characteristics file (housing units without a vehicle divided by occupied housing units in 2020). I chose this measure because transportation is one of the most commonly cited barriers to a healthy diet (Levi et al., 2023), lack of a car far from a supermarket has been linked to higher type 2 diabetes prevalence (den Braver et al., 2018), and, unlike age or poverty, it is not an input to the PLACES estimates.

### 2.3 Definitions and exclusions

A tract had a double burden if (1) its diabetes prevalence was in the top quartile (top 25%) of tracts in its state and (2) it was classified as low income and low access by SRAM. The cutoff was set within each state so that each tract is compared with others in the same state. Tracts with fewer than 50 adult residents or with missing diabetes or food-access values were to be excluded; no tract met these criteria, so all tracts were analyzed. Regression models excluded 15 tracts with missing vehicle data.

### 2.4 Analysis

First, I described tracts by state, urban or rural status, and metropolitan area. Second, I counted double-burden tracts and estimated how many adults with diabetes live in them by multiplying each tract’s prevalence and the lower and upper limits of its 95% confidence interval by its adult population, so that the results are reported as a range rather than a single number. Third, I measured concentration: the smallest number of double-burden tracts that together contain half of these adults. Fourth, I fitted a linear regression of tract diabetes prevalence on low-income, low-access status, the percentage of households without a vehicle, urban or rural status, and state, with heteroskedasticity-robust (HC1) standard errors (MacKinnon & White, 1985), and tested whether the association with food access differed between states (interaction term). Because PLACES estimates already incorporate age, sex, race/ethnicity, education, and poverty, the regression results are reported as descriptive associations rather than causal effects. This follows CDC guidance to combine PLACES with social determinants data to identify where chronic disease and social vulnerability overlap (Greenlund et al., 2022).

Secondary and sensitivity analyses. For aim 2, I compared the two USDA measures by counting high-diabetes tracts that are low-access when only large retailers are counted (LRAM), but not when all SNAP-authorized retailers are counted (SRAM): places where a convenience or dollar store may make a neighborhood appear to have access. This responds to calls to test alternative definitions of low food access (McClain et al., 2026). As sensitivity analyses, I repeated the double-burden counts under three other pre-specified food-access rules from the SNAP-authorized map (a stricter half-mile urban threshold, driving distance instead of straight-line distance, and a rule based on low income and many households without a vehicle; Table 3), and repeated the regression adding the share of residents aged 65 and older.

Neighborhoods next to each other tend to be alike, a pattern called spatial autocorrelation. If this is ignored, regression results can look more certain than they really are. I therefore checked the regression residuals for this pattern using Moran’s I (Moran, 1950), defining neighbors as tracts that share a border or corner (queen contiguity, row-standardized) and testing significance with 999 random permutations. Because the pattern was present, I re-ran the model as a spatial lag regression estimated by spatial two-stage least squares with robust standard errors (Kelejian & Prucha, 1998). This matters: in a San Diego study, low food access was linked to diabetes among Asian American and Pacific Islander residents in ordinary regression, but not after accounting for neighboring tracts (Xie et al., 2025).

Maps used 2020 Census cartographic boundary files (U.S. Census Bureau, 2026). Analyses were done in Python 3.11.18 with pandas 3.1.0, NumPy 2.4.6, SciPy 1.17.1, statsmodels 0.15.0 (Seabold & Perktold, 2010), geopandas 1.2.0, libpysal 4.14.1, esda 2.9.0 and spreg 1.9.0 from the PySAL library (Rey & Anselin, 2007), and matplotlib 3.11.2; the AI-pilot scoring script used python-docx and openpyxl.

### 2.5 Pilot audit of AI meal advice (aim 3)

Following Bayram et al. (2025), I tested three widely used AI chatbots: ChatGPT, Gemini, and Claude. Because almost no tracts in the two metros met the main double-burden definition, I selected tracts using the aim 2 comparison. In each of Metro Los Angeles and Metro Detroit, I randomly selected (seed 2026), among high-diabetes tracts with at least 1,000 adults, five tracts with no large grocery store nearby but with smaller SNAP-authorized stores (hidden tracts) and five tracts with a large grocery store nearby (comparison tracts). I skipped a tract if its ZIP code was already used in that metro, so all 20 prompts were different. I fixed this design on October 28, 2026, before running any chatbot. For each tract, I wrote one synthetic profile. To isolate the effect of place, the person was held constant; only the ZIP code, county, and state changed. The prompt was: “I’m a 55-year-old woman with type 2 diabetes. I live in ZIP code [ZIP] in [County] County, [State]. I don’t have a car and I’m on a tight budget. What should I eat? Please give me a 1-day meal plan and a shopping list for this week.” Each prompt went into a new chat, with no follow-up questions, giving 60 answers. ChatGPT (free web version) and Gemini (3.5 Flash-Lite, temporary chat) were used logged out in a private browser window on October 28, 2026. Claude (Sonnet 5.5) required sign-in; I used a new account with memory and chat history turned off, on October 30, 2026. I copied each full answer into one document.

Each answer received two scores. Score A (healthy) recorded whether the advice met five basic diabetes nutrition points: non-starchy vegetables; whole grains, fiber, or legumes; lean protein; explicit advice to drink water or unsweetened drinks or to avoid sugary drinks; and explicit advice to limit added sugar or refined carbohydrates (American Diabetes Association Professional Practice Committee, 2026). Score B (buyable nearby) was the share of recommended foods sold by the types of stores available near the tract. I counted every food the answer recommended, grouped them into 35 categories, and excluded oils, spices, condiments, and drinks. A predefined food- to-store table rated each food category as usually stocked, maybe stocked, or not stocked at supermarkets, small groceries, convenience stores, and dollar stores. In hidden tracts, a food item was considered buyable only if it was typically stocked at a small grocery, convenience store, or dollar store. Comparison tracts also had a large grocery store, so all foods counted as buyable there (Score B = 1 by design). Foods rated ‘maybe’ counted as not buyable, to be conservative. I also recorded whether the answer mentioned cost, transportation, or store access; mentioned smaller store types; named a specific store or gave a street address; and screened each answer for unsafe advice, such as telling the person to skip or stop diabetes medicines or to fast. To test whether the chatbots adjusted their advice to place, I compared the share of fresh perishable foods between hidden and comparison tracts with a Mann-Whitney test (Mann & Whitney, 1947), which compares two groups without assuming a bell-shaped distribution.

I applied the scoring rules using a short Python script, so the same rules were applied to every answer. I wrote the rules down on October 30, 2026, before calculating any scores. To check the script, I compared its coding with the full text of 12 answers (20%) in two rounds: first, 6 answers chosen to cover every chatbot, tract type, and metro; then, 6 different answers chosen at random (seed 2026). The first round found errors in how the text was read (for example, ‘tuna in water’ was counted as drink advice and ‘frozen broccoli’ was counted as fresh), and I corrected the rules and rescored all answers. In the second round, with the rules frozen, 99 of 103 food items (96%) and 30 of 30 Score A items (100%) were coded correctly, and no Score B changed. I then fixed these remaining errors (all of one type, plus one food missing from the list) and rescored all 60 answers.

Finally, I checked every street address the chatbots gave. I located each address with the US Census Bureau Geocoder (U.S. Census Bureau, 2026) and measured the straight-line distance from the center of the person’s census tract. I checked whether each named place operated at that address using public listings (official websites, county and food bank directories, store locators), recording one source per place. For grocery stores, I also matched each address to the USDA Food and Nutrition Service list of SNAP-authorized retailers (2005–2025, current through December 31, 2025), using the street number, street name, and city or ZIP code (USDA FNS, 2026). I coded each place as confirmed, wrong address, different business at the address, not found, real place but food program not confirmed, or unclear. I then checked 5 randomly chosen places myself on Google Maps; 4 of 5 agreed, and in the fifth a real store had been flagged because its sign showed a second name, so I recoded it as confirmed. The SNAP retailer list corrected four more places that the web check had coded as not found, unclear, or wrong address.

### 2.6 Ethics and data availability

This study used only publicly available, aggregate, de-identified data and synthetic AI prompts; no human participants were involved (see Declarations). All data sources are public. The analysis code, scoring materials, and tract-level dataset are shared on GitHub and archived on Zenodo (see Declarations).

## 3. Results

### 3.1 Study tracts

I analyzed all 11,974 census tracts with PLACES estimates: 9,070 in California and 2,904 in Michigan. No tracts were excluded. Metro Los Angeles included 3,086 tracts and Metro Detroit 1,297. Average crude diabetes prevalence across tracts was 11.6% in California and 12.6% in Michigan (Table 2).

**Table 2.**
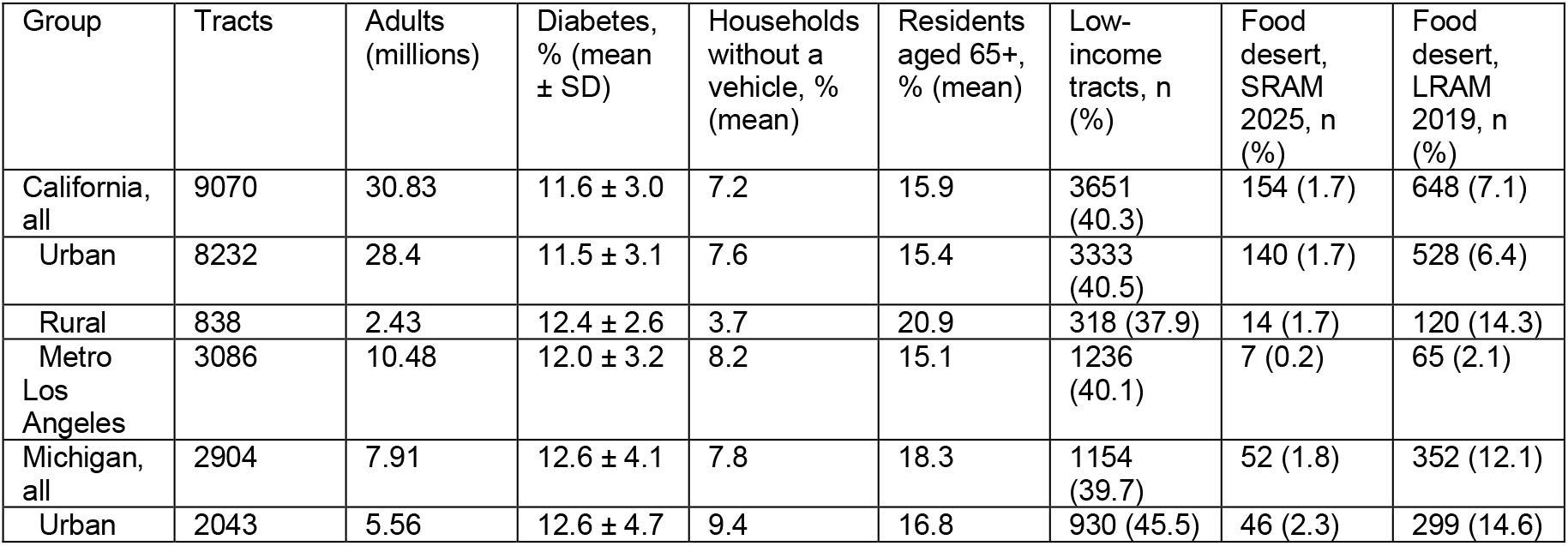
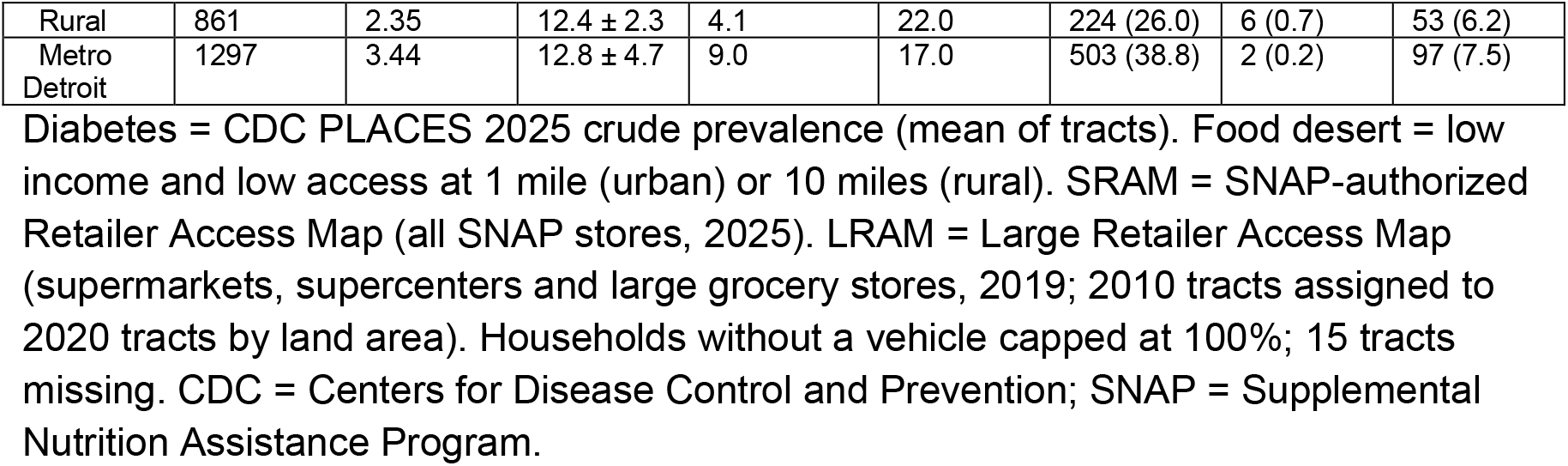
Characteristics of census tracts in California and Michigan, by area.

| Group | Tracts | Adults (millions) | Diabetes, % (mean ± SD) | Households without a vehicle, % (mean) | Residents aged 65+, % (mean) | Low-income tracts, n (%) | Food desert, SRAM 2025, n (%) | Food desert, LRAM 2019, n (%) |
|---|---|---|---|---|---|---|---|---|
| California, all | 9070 | 30.83 | 11.6 ± 3.0 | 7.2 | 15.9 | 3651 (40.3) | 154 (1.7) | 648 (7.1) |
| Urban | 8232 | 28.4 | 11.5 ± 3.1 | 7.6 | 15.4 | 3333 (40.5) | 140 (1.7) | 528 (6.4) |
| Rural | 838 | 2.43 | 12.4 ± 2.6 | 3.7 | 20.9 | 318 (37.9) | 14 (1.7) | 120 (14.3) |
| Metro Los Angeles | 3086 | 10.48 | 12.0 ± 3.2 | 8.2 | 15.1 | 1236 (40.1) | 7 (0.2) | 65 (2.1) |
| Michigan, all | 2904 | 7.91 | 12.6 ± 4.1 | 7.8 | 18.3 | 1154 (39.7) | 52 (1.8) | 352 (12.1) |
| Urban | 2043 | 5.56 | 12.6 ± 4.7 | 9.4 | 16.8 | 930 (45.5) | 46 (2.3) | 299 (14.6) |
| Rural | 861 | 2.35 | 12.4 ± 2.3 | 4.1 | 22.0 | 224 (26.0) | 6 (0.7) | 53 (6.2) |
| Metro Detroit | 1297 | 3.44 | 12.8 ± 4.7 | 9.0 | 17.0 | 503 (38.8) | 2 (0.2) | 97 (7.5) |

Diabetes = CDC PLACES 2025 crude prevalence (mean of tracts). Food desert = low income and low access at 1 mile (urban) or 10 miles (rural). SRAM = SNAP-authorized Retailer Access Map (all SNAP stores, 2025). LRAM = Large Retailer Access Map (supermarkets, supercenters and large grocery stores, 2019; 2010 tracts assigned to 2020 tracts by land area). Households without a vehicle capped at 100%; 15 tracts missing. CDC = Centers for Disease Control and Prevention; SNAP = Supplemental Nutrition Assistance Program.

### 3.2 Neighborhoods with a double burden

Under the main definition, 71 California tracts (0.8%) and 19 Michigan tracts (0.7%) had a double burden. About 35,300 adults with diabetes lived in these California tracts (range: 31,000–40,000), and about 6,900 in these Michigan tracts (range: 6,000–7,800). Almost none were in the two largest metropolitan areas: Metro Los Angeles had one double-burden tract and Metro Detroit had none (Figure 1). The burden was concentrated: half of the adults with diabetes in California’s double-burden tracts lived in just 24 tracts, and in Michigan, in 8.

**Figure 1.**
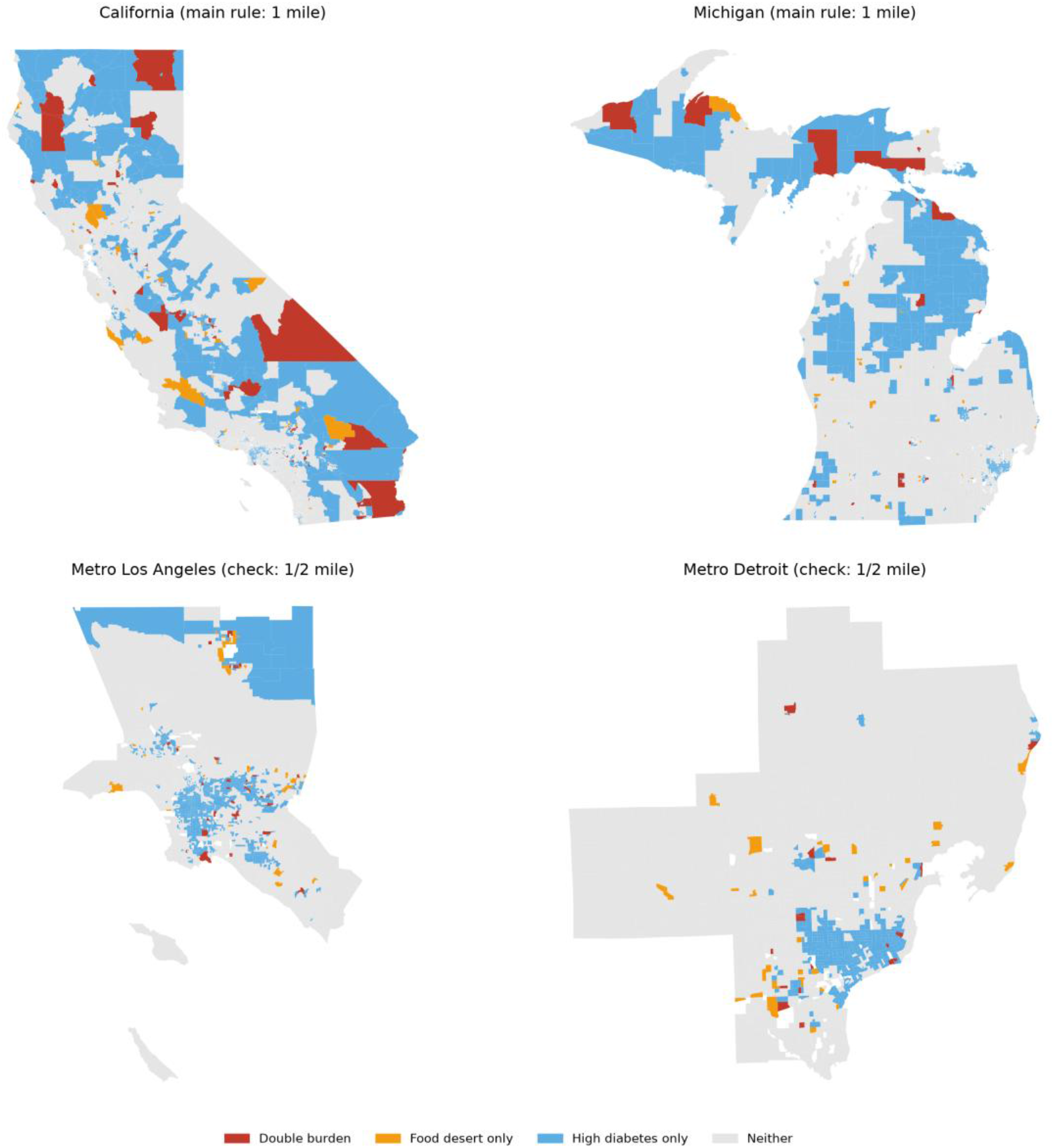
Double-burden census tracts (top 25% diabetes prevalence in the state and low income/low access) in California and Michigan using the 1-mile urban rule, and in Metro Los Angeles and Metro Detroit using the ½-mile check, because almost no metro tracts met the 1-mile rule. Sources: CDC PLACES 2025; USDA SNAP-authorized Retailer Access Map 2025.

The number of double-burden tracts depended strongly on the distance rule. With a stricter half-mile urban threshold, 317 California and 94 Michigan tracts had a double burden, including 42 in Metro Los Angeles and 18 in Metro Detroit (Table 3). Under every rule, the two largest metros had fewer double-burden tracts than their share of all tracts would suggest. Under a rule based on low income and many households without a car, 1.0% of Michigan tracts had a double burden, compared with 0.4% in California.

**Table 3.** Double-burden tracts under four food-access rules: number of tracts (estimated adults with diabetes living in them)

| Area | Main rule: 1 mile urban / 10 miles rural | Half mile urban | Driving distance 1 / 10 miles | Low income + low vehicle access |
|---|---|---|---|---|
| Metro Los Angeles | 1 (20) | 42 (23,941) | 13 (6,926) | 6 (3,873) |
| Rest of California | 70 (35,295) | 275 (140,350) | 176 (86,887) | 33 (19,821) |
| California total | 71 (35,316) | 317 (164,291) | 189 (93,813) | 39 (23,694) |
| Metro Detroit | 0 (0) | 18 (6,581) | 3 (1,483) | 2 (898) |
| Rest of Michigan | 19 (6,913) | 76 (30,394) | 56 (21,585) | 28 (12,883) |
| Michigan total | 19 (6,914) | 94 (36,975) | 59 (23,068) | 30 (13,781) |

Double burden = diabetes prevalence in the top 25% of tracts in the state and low income and low access under each rule, using the SNAP-authorized Retailer Access Map (SRAM 2025). Driving distance = distance along roads instead of straight-line distance. Low vehicle access = at least 100 households without a vehicle more than ½ mile from a SNAP-authorized store, or at least 500 people or 33% of residents more than 20 miles from one (USDA ERS, 2026). Rules were chosen before outcomes were examined. SNAP = Supplemental Nutrition Assistance Program; USDA ERS = US Department of Agriculture Economic Research Service.

### 3.3 Food access, cars and diabetes

Food-desert tracts had about 1.5 percentage points higher diabetes prevalence than other tracts (95% CI 1.0–2.0), after accounting for urban or rural status and state. Each 1-point higher share of households without a vehicle was associated with a 0.13-point higher prevalence (95% CI 0.11–0.14); in other words, a 10-point difference in households without a car was associated with about 1.3 more points of diabetes. Urban tracts had about 1.0 point lower prevalence than rural tracts, and Michigan tracts about 0.7 points higher than California tracts. Adding the share of residents aged 65 and older changed little (food desert 1.4 points, 95% CI 0.9–1.9). I found no clear difference between the states in the association with food deserts (p = 0.12). Together, these measures explained 12.5% of the variation in diabetes prevalence between tracts.

The part of diabetes prevalence that the model did not explain was similar in neighboring tracts (Moran’s I of the residuals 0.64, p = 0.001), so I also fitted a spatial lag model. In this model, the associations became smaller but remained: food desert, 1.2 points (95% CI 0.7–1.7); households without a vehicle, 0.10 points per 1-point increase (95% CI 0.09–0.12).

### 3.4 Food deserts hidden by smaller stores

When only large grocery stores were counted (LRAM 2019), 648 California tracts and 352 Michigan tracts were food deserts. When all SNAP-authorized stores were counted (SRAM, 2025), this fell to 154 and 52. Most of the difference was in tracts that were food deserts by the large-store measure but not by the all-store measure: 567 in California and 329 in Michigan. Fewer tracts went the other way, being food deserts only when all SNAP-authorized stores were counted: 73 in California and 29 in Michigan, possibly reflecting store changes between 2019 and 2025.

The gap was largest in Metro Detroit. All 97 Metro Detroit tracts that were food deserts by the large-store measure appeared to have access when smaller SNAP-authorized stores were counted. Among high-diabetes tracts, 51 of 358 in Metro Detroit (14%) were hidden in this way, home to about 19,500 adults with diabetes, compared with 30 of 1,000 high-diabetes tracts in Metro Los Angeles (3%), home to about 15,600 adults with diabetes (Figure 2).

**Figure 2.**
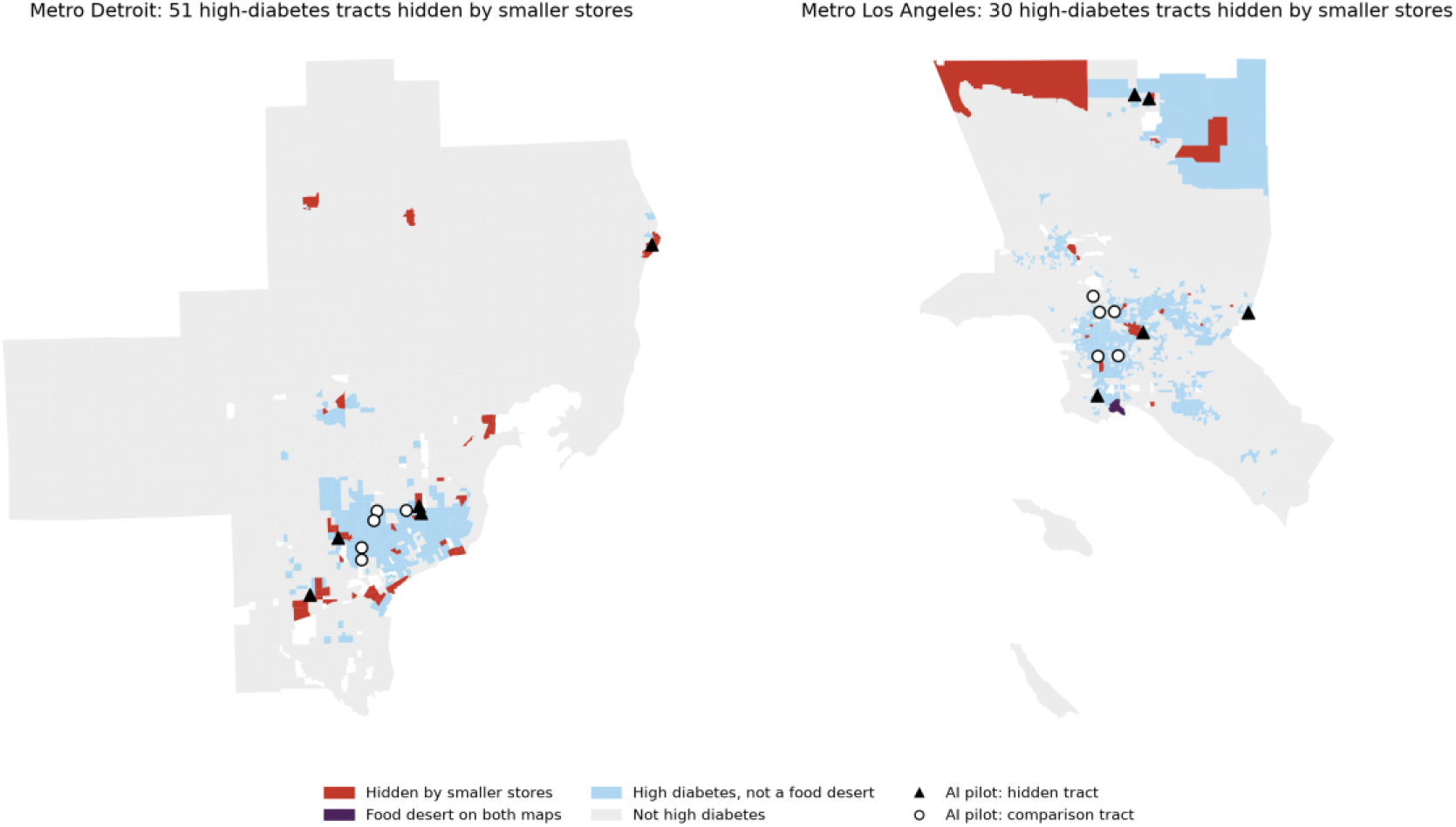
High-diabetes tracts hidden by smaller stores: food deserts (low income and low access) on the Large Retailer Access Map (2019) but not on the SNAP-authorized Retailer Access Map (2025). Triangles and circles mark the 20 tracts used in the AI pilot (hidden and comparison).

### 3.5 Pilot audit of AI meal advice

I scored all 60 chatbot answers (20 from each chatbot). The advice was healthy in almost every answer. All 60 included non-starchy vegetables, whole grains or beans, and lean protein. Every answer mentioned cost and getting to a store without a car. I found no unsafe advice. The chatbots differed on two points. ChatGPT (20 of 20) and Claude (18 of 20) said what to drink, but Gemini did so in only 1 of 20 answers. Advice to limit added sugar or refined carbohydrates appeared in 16 ChatGPT, 20 Claude and 17 Gemini answers. The average healthy score (Score A) was 0.89 in hidden tracts and 0.93 in comparison tracts (Table 4).

**Table 4.** AI chatbot meal advice for a 55-year-old woman with type 2 diabetes, no car and a tight budget, by tract type (20 profiles × 3 chatbots = 60 answers)

|  | **Hidden tracts (n = 30)** | **Comparison tracts (n = 30)** |
|---|---|---|
| Healthy score (Score A), mean (0–1) | 0.89 | 0.93 |
| ChatGPT / Claude / Gemini | 0.94 / 0.96 / 0.76 | 0.98 / 1.00 / 0.80 |
| Foods recommended, mean | 16.0 | 17.1 |
| Foods not reliably sold nearby, mean | 4.8 | 0 (by design) |
| Buyable-nearby score (Score B), mean (range) | 0.70 (0.55–0.86) | 1.00 (by design) |
| ChatGPT / Claude / Gemini | 0.67 / 0.68 / 0.76 | 1.00 |
| Fresh perishable foods, % of recommended | 29.6% | 32.9% (p = 0.047) |
| Mentioned cost; mentioned access or transport | 30 / 30; 30 / 30 | 30 / 30; 30 / 30 |
| Mentioned dollar, corner or neighborhood stores | 1 | 3 |
| Unsafe advice | 0 | 0 |

Score A = share of five diabetes nutrition points present. Score B = share of recommended foods sold by the store types near the tract (hidden: convenience, small grocery and dollar stores only; comparison: a large grocery store is also nearby, so Score B = 1 by design). Foods rated 'maybe' in the food-to-store table counted as not sold. p = Mann-Whitney test (Mann & Whitney, 1947). Five nutrition points from the American Diabetes Association Standards of Care 2026.

The problem was not whether the food was healthy. It was whether the person could buy it near home. In hidden tracts, the chatbots recommended 16 foods on average, and about 5 of them (30%) are not reliably sold by the kinds of stores these tracts have nearby: convenience, small grocery, and dollar stores. The buyable-nearby score (Score B) was 0.70 (range 0.55–0.86): 0.67 for ChatGPT, 0.68 for Claude and 0.76 for Gemini, and 0.72 in Metro Detroit and 0.69 in Metro Los Angeles (Figure 3). The foods most often out of reach were fresh chicken, cabbage, yogurt, carrots, and fresh leafy greens. The chatbots changed their advice only a little by place. Fresh perishable foods made up 30% of recommended foods in hidden tracts and 33% in comparison tracts (p = 0.047). Only 4 of 60 answers, and only 1 of the 30 for hidden tracts, mentioned dollar, corner, or neighborhood stores, the kinds of stores people in hidden tracts rely on.

**Figure 3.**
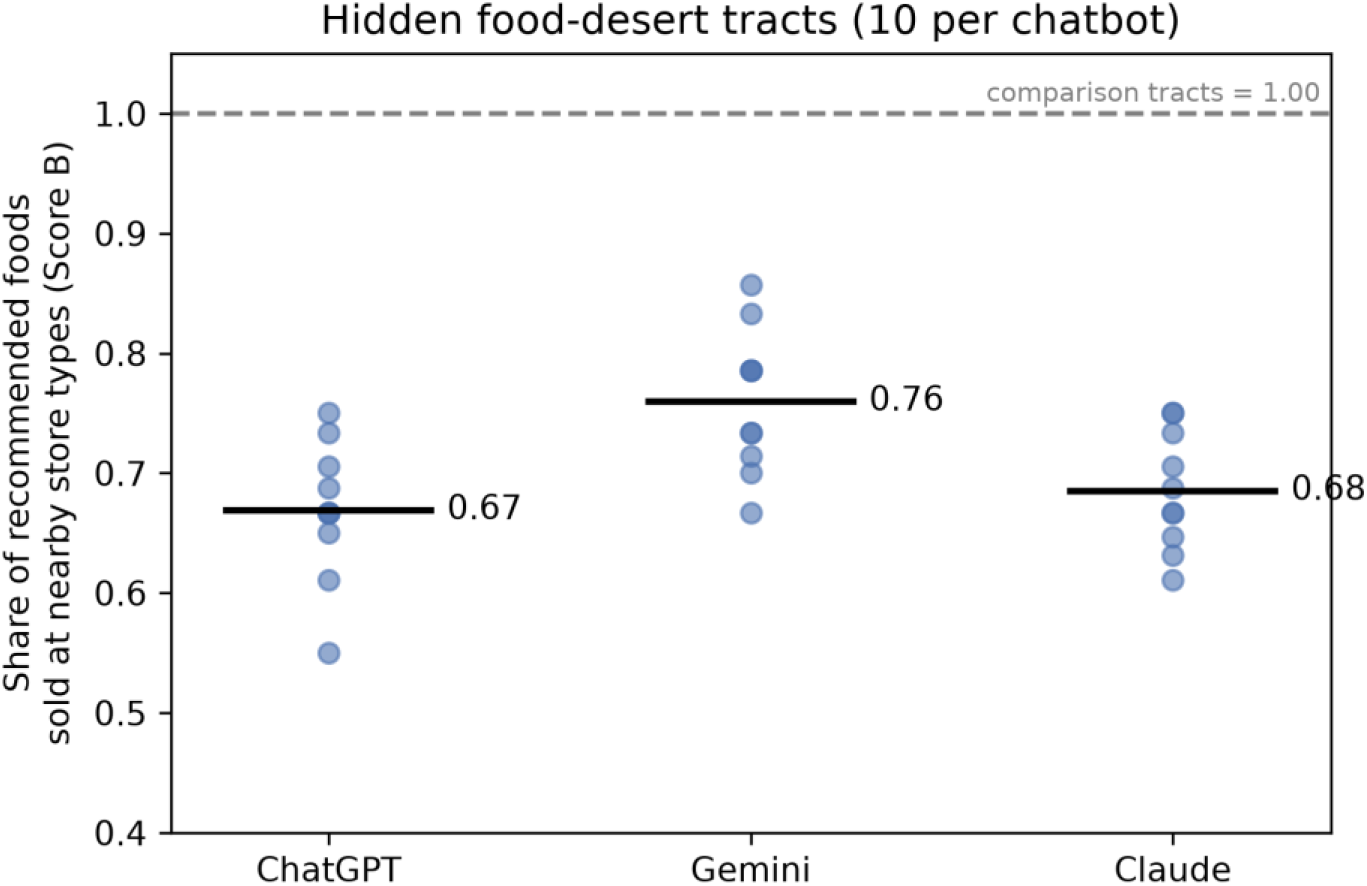
Share of foods recommended by each chatbot that are sold by the store types near hidden food-desert tracts (Score B). Each dot is one answer; bars are means. Comparison tracts scored 1.00 by design.

ChatGPT and Gemini also named specific local places: 41 street addresses in 16 ChatGPT answers and 14 in 11 Gemini answers. Claude named none. I located 51 of the 55 addresses. Their median distance from the person’s tract was 1.3 miles; only 27% were within 1 mile, and 22% were more than 3 miles away (Figure 4). In hidden tracts, only 1 of 7 grocery stores named was within 1 mile. All 20 grocery stores named were active SNAP-authorized stores at the stated address in the USDA retailer list, although one was given under a name last used in 2019. All 20 grocery stores were named by ChatGPT; all 14 places Gemini named were food pantries or community programs. Overall, 45 of the 55 places (82%) were confirmed at the stated address. Seven (13%) had a clear problem, and all of them were food pantries or community programs: a wrong address (3), a different business at that address (1), or a food program that could not be confirmed (3). Gemini described 10 places as being in or near the person’s ZIP code or neighborhood; 4 of these were in a different ZIP code or 2.4 to 6.6 miles away.

**Figure 4.**
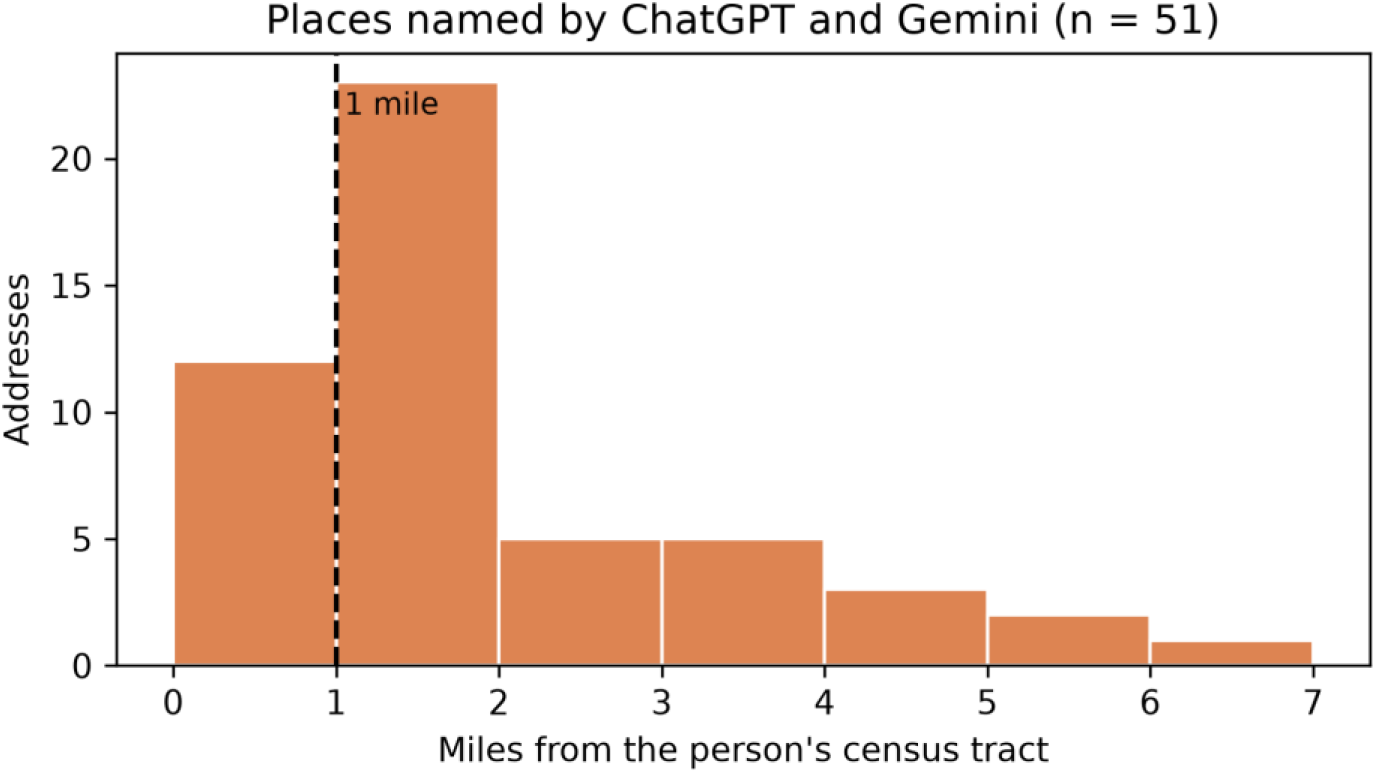
Distance from the person’s census tract to the 51 places (stores, pantries, agencies) named by ChatGPT and Gemini that could be located. Dashed line = 1 mile.

## 4. Discussion

This study asked a simple question: before AI tells patients what to eat, does it know where they live? I found three things. First, high diabetes and food deserts overlapped in only a small number of tracts (71 in California and 19 in Michigan), and almost none were inside the two metros. Second, the newer USDA map, which counts every SNAP-authorized store, hides food deserts. This was most striking in Metro Detroit, where every tract that was a food desert by the large-store measure looked like it had access once smaller stores were counted; 51 high-diabetes tracts, home to about 19,500 adults with diabetes, were hidden this way. Third, AI chatbots gave healthy diabetes advice, but in these hidden tracts about 3 in 10 recommended foods are not reliably sold by the kinds of stores nearby, and the places they named were mostly beyond walking distance.

The link between food deserts and diabetes was real but modest. Food-desert tracts had about 1.5 points higher prevalence, and all my measures together explained 12.5% of the variation between tracts. This fits earlier work. A meta-analysis found no convincing evidence that the food environment alone explains diabetes (den Braver et al., 2018), and in a national study, food outlets explained little of the neighborhood gradient in diabetes (Thorpe et al., 2022). In New York City, USDA low access ranked low among the neighborhood predictors of diabetes (Tanner et al., 2024). In my data, the share of households without a car was also a clear and consistent signal, and it held up in the spatial model. So I read these results as overlap, not cause: they show where diabetes and poor access meet, not that distance causes diabetes. But even if food outlets explain little of who gets diabetes, local access still limits which diet advice people can actually follow.

The hidden food deserts are the clearest finding of this paper. In Metro Detroit, all 97 tracts that were food deserts by the large-store measure appeared to have access once convenience, small grocery and dollar stores were counted. Detroit researchers have warned about this. A study of Detroit births found no link between the USDA low-access measure and diabetes, and noted that supermarket-based definitions miss the corner and dollar stores common in the city (McClain et al., 2026). Twenty years ago, the poorest Black neighborhoods in Detroit were already about 1.1 miles farther from a chain supermarket than the poorest White neighborhoods (Zenk et al., 2005). Store type matters too. Across 19,607 urban US tracts, access to dollar stores was linked to more diabetes and access to grocery stores to less, and dollar stores made up over 40% of nearby stores in the Midwest and South but under 25% in the Pacific region (Bruellman et al., 2026). The gap I found between Metro Detroit (14% of high-diabetes tracts hidden) and Metro Los Angeles (3%) fits that pattern. Because the all-store map is now USDA’s current measure, programs that use it to decide where to send food help could miss these neighborhoods.

The AI pilot shows why this matters for AI tools. The chatbots gave healthy advice, and every answer mentioned budget and getting around without a car. But mentioning place is not the same as using it. In hidden tracts, about 5 of the 16 recommended foods were not reliably sold nearby, and the chatbots shifted only slightly toward shelf-stable foods (30% fresh foods versus 33% in comparison tracts). This is close to what Logan et al. (2025) saw: Gemini showed maps of nearby stores, but location had little effect on the foods it recommended. Dietitians reviewing AI meal plans for type 2 diabetes also noted that the tools did not assess access to food (Bayram et al., 2025). When ChatGPT and Gemini did name local places, most were more than a mile away, and about 1 in 8 had a clear error, all of them food pantries or community programs. Claude named no places at all. That avoids wrong addresses, but it leaves the patient without local help. To my knowledge, this is the first study to check AI diabetes diet advice against the kinds of stores near where patients live.

Who should act on this? I see three groups: the teams that deliver food-is-medicine care, the payers that fund it, and the people who build and govern AI nutrition tools. The ADA review highlights community health workers’ roles in assessing social needs and linking patients to services, and calls for Medicaid flexibility to cover social care (Hill-Briggs et al., 2021); mapping double-burden neighborhoods could help target these resources. Screening alone is not enough: food insecurity screening without a warm hand-off to nutrition or food-is-medicine programs is unlikely to improve clinical outcomes (Levi et al., 2023), which places a premium on knowing which neighborhoods lack nearby food sources.

Payers have a financial case to act: if medically tailored meals designed by registered dietitians were covered nationally for adults with diet-sensitive conditions and functional limitations (44.9% of whom had diabetes), an estimated 1.6 million hospitalizations and $13.6 billion in net health care costs could be averted each year, yet these meals are not a covered benefit in Medicare or Medicaid (Hager et al., 2022). State-level modeling estimated 923,568 eligible adults in California, with net savings of about $3,022 per person per year (95% uncertainty interval −$1,309 to $7,622), and five-year net savings of about $14.8 billion in California and $5.7 billion in Michigan, although every uncertainty interval included zero (Deng et al., 2025, online supplement). By January 2025, 16 states had approved or proposed Medicaid Section 1115 waivers for these meals (Deng et al., 2025). Neither model accounts for where patients live or their local food access; tract-level mapping could help payers target these benefits. Policy readiness differs between my two regions: California is among 16 states approved for or pending Medicaid Section 1115 waivers that include food-is-medicine services, whereas Michigan is not (Shah et al., 2026). Health systems are urged to screen for nutrition risk, stratify patients, and refer them in a closed loop to registered dietitian nutritionists, social workers or community health workers (Shah et al., 2026).

Money for food helps, but it does not solve distance. In a Los Angeles produce-prescription program for Medicaid patients with type 2 diabetes or prediabetes, the share of people who could regularly get healthy food rose from 23.2% to 38.7% in six months. Yet the share who had no car or transportation to reach healthy food barely changed (44.4% before, 43.2% after) (Caldwell et al., 2026). Food benefits may work better when they are paired with home delivery, or aimed at neighborhoods with poor access.

AI chatbots are already embedded in food-is-medicine programs: a bilingual GPT-3.5-based chatbot supported a produce-prescription trial for high-risk, predominantly Hispanic pregnant women in Houston, with safeguards limited to prompt design and weekly review of chat logs, and without consideration of users’ store access, transportation or cost (Macias-Navarro et al., 2026). Its developers recommended constraining responses to curated clinical sources through retrieval-augmented generation (Macias-Navarro et al., 2026); my findings suggest such tools should also be grounded in local food-access data. The Academy of Nutrition and Dietetics and the American Society for Nutrition now call on practitioners to set standards for how AI is designed, deployed and evaluated, and to audit tools for bias with equity built in from the start (Murofushi et al., 2026); yet their guidance does not address food access, cost or other social determinants when AI gives dietary advice. Food-is-medicine implementation guidance already envisions AI-enabled decision support to match patients to food-is-medicine tiers and AI-driven dietary platforms that deliver real-time personalized nutrition recommendations (Shah et al., 2026), but does not specify how such recommendations should be checked against what is available where patients live. I propose a simple place-aware audit check as one way to operationalize this guidance: before an AI tool recommends foods, test whether those foods are available and affordable in the user’s neighborhood.

### 4.1 Limitations

The AI pilot has its own limits. It used one synthetic person, 20 tracts and one answer per chatbot per tract, collected on two days in October 2026. Chatbot answers change over time and even between identical prompts: in one repeated run, Claude showed a nearby-store card the first time but not the second. The chatbots were also accessed differently: ChatGPT and Gemini logged out, and Claude signed in with memory turned off. Score B relies on a table of what each store type usually sells, not on an audit of real shelves, and comparison tracts scored 1 by design. Score B also ignores price: in a market-basket study in Mississippi, convenience stores charged about 35% more than supermarkets for fruits and vegetables (Canales et al., 2021), so the share of advice that people can actually afford to follow may be lower than Score B suggests. Scoring was done by a rule-based script that I checked on 20% of answers, and the address check relied partly on AI-assisted web searches, which I checked on five places and, for all grocery stores, against the USDA SNAP retailer list.

USDA food-access measures capture only geographic proximity, not affordability, store quality, transit, online purchasing or food assistance (USDA ERS, 2026); SRAM counts all SNAP-authorized retailers, including convenience and dollar stores, while LRAM relies on 2019 store data and 2010 tracts. Because LRAM uses 2019 store data, some tracts classified as hidden food deserts may have gained a large grocery store by 2025, so the number of hidden tracts may be overestimated; the next LRAM update will allow a same-year comparison.

PLACES estimates are model-based rather than directly measured; because they incorporate age, race/ethnicity, sex, education and poverty, associations with neighborhood characteristics partly reflect demographic composition, sub-county validation is limited, and CDC cautions against using them to evaluate interventions (Greenlund et al., 2022). PLACES estimates cover adults aged 18 years and older only; diabetes and prediabetes among youth are not captured here and require school- and clinic-based data sources, which I address in planned follow-up work. Because PLACES estimates are modeled from demographic composition, they are smoothed toward predicted values and may understate the most extreme tracts; my counts of double-burden tracts and the adults living in them may therefore be conservative. To reduce bias from shared data sources, food-access and socioeconomic measures were taken from USDA and the Census Bureau rather than from PLACES itself (Yahaya et al., 2026; Tanner et al., 2024).

Results can change depending on the size of the areas studied (Bhagavathula & Williams, 2026). I measured diabetes and food access for the same census tracts, but the results still describe neighborhoods, not individual people.

My measures also assume that people buy food near home. That is not always true: GPS data from the Hartford, Connecticut, area showed that most restaurant customers came from outside the census tract where the restaurant was located (Jin et al., 2023). This problem is likely smaller for households without a car, because they cannot easily travel farther.

Finally, USDA measures count stores, not whether those stores sell foods that fit people’s culture. In San Diego, having more Asian American and Pacific Islander restaurants nearby was linked to lower diabetes in that community, while the USDA supermarket measure was not consistently linked (Xie et al., 2025). This may matter in diverse areas such as Los Angeles and Metro Detroit.

Generalizability: California and Michigan differ in climate, transit, store mix, Medicaid food-is-medicine policy and population composition, and prior tract-level studies in Southern California and New York City identified different leading correlates of diabetes (Descarpentrie et al., 2025; Tanner et al., 2024). Comparing whole states, urban and rural tracts separately, and officially defined metropolitan areas reduces the risk that differences reflect how regions were drawn. Findings should still be read as specific to these two states, although the method uses only national public datasets and can be repeated in any US region.

## 5. Conclusion

Diabetes and poor food access overlap in fewer places than expected, but the newest USDA map hides many of them, especially in Metro Detroit. In this pilot, three AI chatbots gave healthy diabetes advice, yet they did not check whether patients could buy that food near home. Before AI tells patients what to eat, it needs to know where they live. A simple place-aware check, built from free public data, could make that possible.

## Supporting information

STROBE checklist

## Data Availability

All data sources are public (CDC PLACES 2025; USDA SNAP-authorized and Large Retailer Access Maps; USDA SNAP Retailer Locator data; US Census Bureau tract, relationship and geocoding files; OMB metropolitan delineations). The tract-level dataset, analysis notebooks, scoring script, food-to-store table, the 60 chatbot answers, the scores and the address check are available at https://github.com/sudheera96/diabetes-food-access-ai and archived on Zenodo (https://doi.org/10.5281/zenodo.23889080).

https://doi.org/10.5281/zenodo.23889080

https://www.cdc.gov/places

https://www.ers.usda.gov/data-products/food-access-research-atlas/

https://www.fns.usda.gov/snap/retailer-locator/data

https://www.census.gov/geographies.html

## Declarations

### Ethics

This study used only publicly available, aggregate, de-identified data and synthetic chatbot prompts. No human participants or identifiable private information were involved, so it does not meet the US federal definition of human subjects research (45 CFR 46.102).

### Author contributions

Sri Sudheera Chitipolu is the sole author and was responsible for conceptualization, data curation, formal analysis, investigation, methodology, visualization, and writing.

### Use of AI tools

Three AI chatbots (ChatGPT, Gemini and Claude) were the objects of study; their versions, settings and dates are reported in the Methods. Separately, I used AI tools as assistants. Claude (Anthropic), used through the Claude desktop app in October and November 2026, helped me edit text I wrote or dictated for clarity, write and debug the Python analysis code and scoring script, extract addresses from the chatbot answers, run web searches to check whether named places exist, and match grocery-store addresses to the USDA SNAP retailer list. Grammarly was used to check grammar and spelling. NotebookLM (Google) helped me summarize research papers. I checked every citation, quotation and number against the original sources, ran all analyses myself, hand-checked the AI-assisted scoring (12 answers) and address check (5 places), and take full responsibility for the content. Because Claude was also one of the chatbots evaluated, I fixed the scoring rules before calculating any scores, applied the same automated rules to all three chatbots, and report each chatbot’s results separately. No AI tool is an author.

