## Supplementary material for "Before AI tells patients what to eat, it needs to know where they live: a cross-sectional ecological study of diabetes and food access in California and Michigan census tracts, with a spotlight on Metro Detroit and Metro Los Angeles": STROBE checklist

### STROBE checklist (cross-sectional studies)

Manuscript: "Before AI tells patients what to eat, it needs to know where they live: a cross-sectional ecological study of diabetes and food access in California and Michigan census tracts, with a spotlight on Metro Detroit and Metro Los Angeles." Author: Sri Sudheera Chitipolu. Page numbers refer to the submitted manuscript PDF. Note: this is an ecological study; the unit of analysis is the census tract, not individual participants.

| Section/Topic | Item | Recommendation (abbreviated) | Reported on page |
|---|---|---|---|
| Title and abstract | 1 | (a) Study design in title or abstract | 1 (title: "cross-sectional ecological study") |
| | | (b) Informative and balanced summary in abstract | 1 |
| Introduction: Background/rationale | 2 | Scientific background and rationale | 1–3 |
| Objectives | 3 | Specific objectives, prespecified hypotheses | 3 (three aims) |
| Methods: Study design | 4 | Key elements of study design early in paper | 3 (2.1) |
| Setting | 5 | Setting, locations, relevant dates | 3–4 (2.1, 2.2); chatbot data collection dates 6–7 (2.5) |
| Participants | 6 | (a) Eligibility criteria, sources and selection | 5 (2.3: census tracts with PLACES estimates; exclusion rules); AI-pilot tract selection 6 (2.5) |
| Variables | 7 | Define outcomes, exposures, predictors, confounders, effect modifiers | 4–6 (2.2–2.4); Table 1, p. 4 |
| Data sources/measurement | 8 | Sources and methods of measurement for each variable | 4–5 (2.2, Table 1); scoring and address check 7–8 (2.5) |
| Bias | 9 | Efforts to address potential sources of bias | 5–6 (independent data sources, pre-specified rules, spatial model); 7 (locked scoring rules, spot checks); limitations 16–17 |
| Study size | 10 | How study size was arrived at | 3, 8 (all 11,974 tracts with PLACES estimates; 20 pilot tracts × 3 chatbots = 60 answers, 6) |
| Quantitative variables | 11 | How quantitative variables were handled; groupings | 5 (top-quartile diabetes; low-income/low-access flags; % no vehicle; % aged 65+) |
| Statistical methods | 12 | (a) All statistical methods, incl. confounding control | 5–6 (2.4) |
| | | (b) Subgroups and interactions | 5–6 (state, urban/rural, metro; state interaction term) |
| | | (c) Missing data | 5 (15 tracts with missing vehicle data excluded from regression) |

| | | | |
|---|---|---|---|
| | | (d) Sampling strategy | Not applicable (all tracts analyzed; PLACES are model-based estimates, 4) |
| | | (e) Sensitivity analyses | 6 (three alternative food-access rules; age 65+ model; spatial lag model) |
| Results: Participants | 13* | (a) Numbers at each stage | 8 (3.1: 11,974 tracts; none excluded); pilot 12 (60 answers, 55 addresses, 51 located) |
| | | (b) Reasons for non-participation | Not applicable (no tracts excluded, 8) |
| | | (c) Flow diagram | Not applicable |
| Descriptive data | 14* | (a) Characteristics of study participants | 8–9 (Table 2) |
| | | (b) Missing data for each variable | 8 (Table 2 note: 15 tracts missing vehicle data) |
| Outcome data | 15* | Numbers of outcome events or summary measures | 9 (double-burden counts, Table 3); 11 (hidden food deserts); 12–13 (AI pilot, Table 4) |
| Main results | 16 | (a) Unadjusted and adjusted estimates with precision | 10 (3.3, 95% CIs; spatial model) |
| | | (b) Category boundaries for continuous variables | 5 (top 25% diabetes prevalence within state) |
| | | (c) Relative to absolute risk | Not applicable |
| Other analyses | 17 | Subgroups, interactions, sensitivity analyses | 9 (Table 3, four access rules); 10 (age 65+, state interaction, spatial model); 11 (hidden food deserts); 12–13 (AI pilot) |
| Discussion: Key results | 18 | Summarise key results with reference to objectives | 14 |
| Limitations | 19 | Limitations, sources of bias or imprecision | 16–17 (4.1) |
| Interpretation | 20 | Cautious overall interpretation | 14–16 |
| Generalisability | 21 | External validity | 17 |
| Other information: Funding | 22 | Funding source and role of funders | 18 (Declarations) |

*Applies to the census tracts analyzed (ecological units). STROBE checklist source: von Elm E, et al. The Strengthening the Reporting of Observational Studies in Epidemiology (STROBE) statement. https://www.strobe-statement.org
